# Unguided Web-Based Brief Intervention with Genetic Risk Education to Reduce Unhealthy Alcohol Consumption: Protocol for a Randomized Controlled Trial

**DOI:** 10.1101/2025.07.10.25331236

**Authors:** Yuki Kono, Mai Tanaka Sahker, Shino Kikuchi, Yan Luo, Masatsugu Sakata, Ryuhei So, Tamara L Wall, Toshi A Furukawa, Ethan Sahker

## Abstract

**Background:** Alcohol use is one of the largest worldwide contributors to morbidity and mortality, and a major risk factor for esophageal cancer. Reducing or stopping alcohol use results in a significant esophageal cancer risk reduction. A causal factor in alcohol-related esophageal cancer is exposure to acetaldehyde, an alcohol metabolite and established carcinogen. Japanese people have a high proportion of the aldehyde dehydrogenase 2*2 (*ALDH2*2*) allele, which causes a deficiency in ALDH2 enzyme activity and increases exposure to acetaldehyde, elevating their risks of esophageal cancer. Personalized health information about such risks can be important in developing motivation to moderate alcohol consumption. Fortunately, brief intervention (BI) for unhealthy alcohol use is an effective behavioral intervention that can capitalize on this motivation source but is underused in Japan.

**Objectives:** The proposed project will test the efficacy of an unguided web-based BI using genetic cancer risk education to reduce alcohol consumption in a randomized controlled trial.

**Methods:** Participants will be recruited online and screened for moderate alcohol use and probable *ALDH2*2* allele. Included participants will be randomized to an experimental BI condition or sham educational control. The experimental condition will receive an unguided web-based brief video intervention. The intervention will educate participants about their probable genetic risk, how consuming alcohol significantly increases risk of esophageal cancers, and the benefits of reducing or stopping alcohol use. The primary outcome is mean endpoint past 4-week alcohol quantity at 3-months post-randomization. Additionally, secondary main effects will be investigated with alcohol use in grams, severity, motivation to change, health knowledge retention, participant satisfaction, and quality of life. Assessments will be collected at baseline, 1- , 2-, and 3-months post-randomization through a web portal.

**Results:** The present study was funded in April 2024. Data collection is projected to occur August 2025 – August 2026 through a Japanese research panel company. Currently, no data has been collected.

**Conclusions:** This intervention is expected to reduce unhealthy alcohol use at a low cost of implementation by using personalized health information as a motivational factor in a general population. Additional group differences are expected to be observed in secondary alcohol use outcomes.

**Trial Registration:** This trial has been registered in University Hospital Medical Information Network Clinical Trials Registry (UMIN000058012).

## BACKGROUND

Alcohol use is a leading causes of disease burden worldwide [1] and a major risk factor for cancer [2]. Of all alcohol-related forms, esophageal cancer has the highest alcohol-attributable fraction [3]. That is, the greatest proportion of this type of cancer would be prevented if no alcohol was consumed. In Japan, the estimated disease burden due to alcohol-related esophageal cancer is large with an estimated 5,279 cancer deaths and 102,988 disability adjusted life years lost per year [4]. Acetaldehyde, an alcohol metabolite and established carcinogen, is a causal factor in alcohol-related esophageal cancer [5]. Alcohol metabolizing genetic variations are known to influence risk for esophageal cancer [6]. The aldehyde dehydrogenase 2 (*ALDH2*) gene encodes the primary enzyme involved in the metabolic removal of acetaldehyde. The *ALDH2* genotype shows the strongest association with esophageal cancer. The variant *ALDH2*2* allele encodes a less active version of the ALDH2 enzyme, resulting in impaired acetaldehyde metabolism [7]. Thus, those with an *ALDH2*2* allele are exposed to higher levels of acetaldehyde during alcohol consumption, but can still consume alcohol [8]. Although the *ALDH2*2* variant has an overall protective association with alcohol consumption and alcohol use disorder [9], a significant proportion of people with *ALDH2*2* are moderate or heavy drinkers [6].

Those with *ALDH2*2* show an increasing risk for esophageal cancer as alcohol consumption increases in a clear dose-response relationship [6], which is substantially greater than those without the *ALDH2*2* allele. Esophageal cancer risk outcomes associated with alcohol use in people with the ALDH2*2 allele is highly varied with relative risks (RR) ranging from 1.5 times the risk for all drinkers to 89 times the risk for heavy drinkers [10–13]. Summary literature has confirmed the exponential dose response with increasing alcohol consumption [10].

Conservative risk estimates are still substantial, with 2 times greater risks for light drinkers with the *ALDH2*2* allele, 2.4 times for moderate drinkers, and 4.4 times for heavy drinkers. The Japanese population has a high occurrence of the *ALDH2*2* allele, estimated to be between 41-52% [14]. The presence of an *ALDH2*2* allele presents through “Asian flush,” a reddening of the face caused by accumulation of acetaldehyde from alcohol metabolic functions [14].

Importantly, identifying people at risk and developing strategies to stop or moderate alcohol use, results in a significant reversal in esophageal cancer risk [15]. Nonetheless, this alcohol-related risk remains under-recognized in Japan and interventions are needed to improve health outcomes [6].

The availability of effective brief interventions (BIs) for alcohol use [16] present an opportunity to direct primary prevention efforts toward those at increased genetic risk for alcohol-related cancers. BIs range in application, but are generally delivered in 1 to 5 individual sessions lasting between 5 to 60 minutes each and is indicated for low to moderate drinking [17]. BI is based in social, cognitive, and behavioral approaches of psychotherapy [16]. The overarching components of BI include (a) feedback about alcohol use and harm, (b) risks of continued use, (c) benefits of reducing use, (d) advice about how to reduce use, (e) motivational enhancement, and (f) how to develop a plan to reduce use [16]. These components have been administered through the transtheoretical model of change [18] using the FRAMES approach: Feedback, Responsibility to change, Advice, Menu of treatment options, Empathy, and supporting of Self-efficacy [16,19–21].

Effective BI are attractive approaches for behavioral healthcare interventions due to low costs and scalability. However, extant BIs using healthcare risk as motivation to change have yet to test cost-effective and scalable interventions. The efficacy of a non-invasive unguided web-based delivery of BI using personalized health risks as a motivational intervention in Japan remains untested. There is nascent evidence of BI for alcohol moderation in Asian populations, but they remain invasive and costly. A trial with Japanese university students reported that BI using *ALDH2*2* allele associated cancer risks significantly reduced alcohol quantity and severity, while also improving motivation to change, compared to education only control [22]. However, this trial used invasive blood draws for genotyping and resource consuming in person BI, increasing costs and reducing ease of implementation. Another pilot study conducted in the U.S. with Asian American university students used a web-based BI approach and included *ALDH2*-specific genetic alcohol-related cancers risk information. This study showed evidence for the feasibility, acceptability, and efficacy for reducing alcohol use in individuals with *ALDH2*2* in the U.S. [23]. While this study capitalized on web-based BI, they also used invasive blood draws for genotyping, increasing costs, reducing ease of implementation, and requiring human contact. An unguided approach to BI with simple and non-invasive *ALDH2* screening represents a much more cost-effective delivery format that can increase identification and intervention among non-treatment-seeking populations [24,25]. Yet, such a non-invasive unguided web-based delivery of BI focused on personalized health risks has yet to be proven for alcohol moderation.

The present study will investigate the efficacy of a non-invasive and unguided web-based BI using genetic risk education (BIGRE) to moderate alcohol consumption in a Japanese general population. This approach presents less costly and more scalable intervention approaches, reducing healthcare system burden. Consistent with personalized medicine and public health genetics perspectives [26], the present parallel group randomized controlled trial will screen for alcohol use and probable *ALDH2*\*2 genotypes. We will randomize participants to receive either the BIGRE intervention or a sham quality of life information control (QoL Control).

Hypotheses:

1. Participants in the BIGRE intervention group will report fewer past-month standard alcohol drinks than the sham control group at 3-months post randomization.
2. Other between-group differences will be observed in secondary outcomes.
3. Results will help inform future iterations of unguided web-based BI interventions in terms of development and feasibility.

## METHODS

The present study was approved by the Kyoto University Graduate School of Medicine Ethics Committee (C1711-1) and has been registered in University Hospital Medical Information Network Clinical Trials Registry (UMIN000058012). This protocol follows the SPIRIT reporting guidelines [27].

### Participants

Participants will be recruited from a Japanese adult general population through a research survey panel of nearly 29 million Japanese members via NEO Marketing Inc. Academic Research Services. Based on established allele frequencies in the Japanese population [14], we anticipate that 41-52% of people will possess probable *ALDH2*2*.

#### Inclusion Criteria

1. 20 to 64 years of age
2. At least 4 drinks per week on average
3. Positive screening for probable *ALDH2*\*2 allele

#### Exclusion Criteria

1. Current alcohol use treatment
2. Lifetime cancer diagnosis
3. Pregnant

#### Study Setting

The study setting will consist of an internet panel sample in Japan.

### Procedure

Figure 1 shows the proposed timeline. Recruitment will be conducted by Neo Marketing, a marketing company specialized in academic research. Neo Marketing will invite people from their survey panel to participate in a study providing the opportunity to learn about health improvement. Participants will complete screening based on alcohol use quantity, probable *ALDH2*2* genotype, cancer history, current SUD treatment, pregnancy status, and age. Outcome assessments will be conducted at baseline then participants will be randomized to intervention groups. After interventions, participants will be assessed on health risk knowledge retention and program satisfaction. Post-randomization follow-up assessments will be collected at 1-, 2-, and 3-months. At follow-up, participants will be contacted through email or LINE and asked to complete questionnaires. Interventions are provided immediately after randomization and allocation.

**Figure 1.**
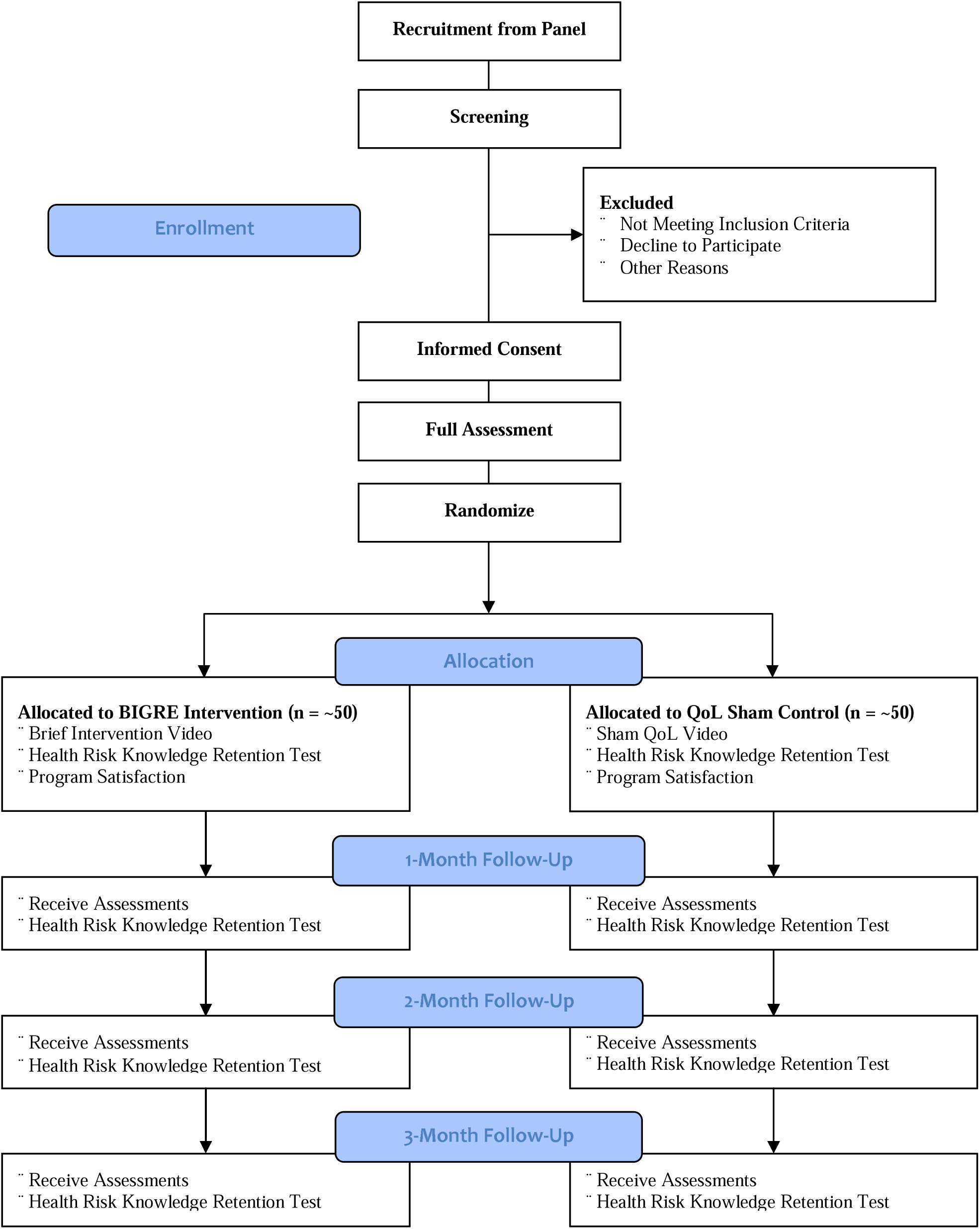
Web-Based Screening and Brief Intervention with Genetic Cancer Risk Education to Reduce Alcohol Consumption.

#### Screening

Alcohol use quantity will be screened by asking, “Using the standard drinks conversion chart, how many drinks do you have in a typical week?” We will prompt participants with the definition conversion chart (Figure 2) to determine standard drinks: 250ml beer (∼5% alcohol), 90ml wine/sake (∼15% alcohol), 45ml spirits (40% alcohol) [28]. Based on power calculations (see below), a minimum of 14 standard drinks in the past 4 weeks is required to detect a change. Thus, participants will be included if they report drinking 4 or more standard drinks in an average week.

**Figure 2.**
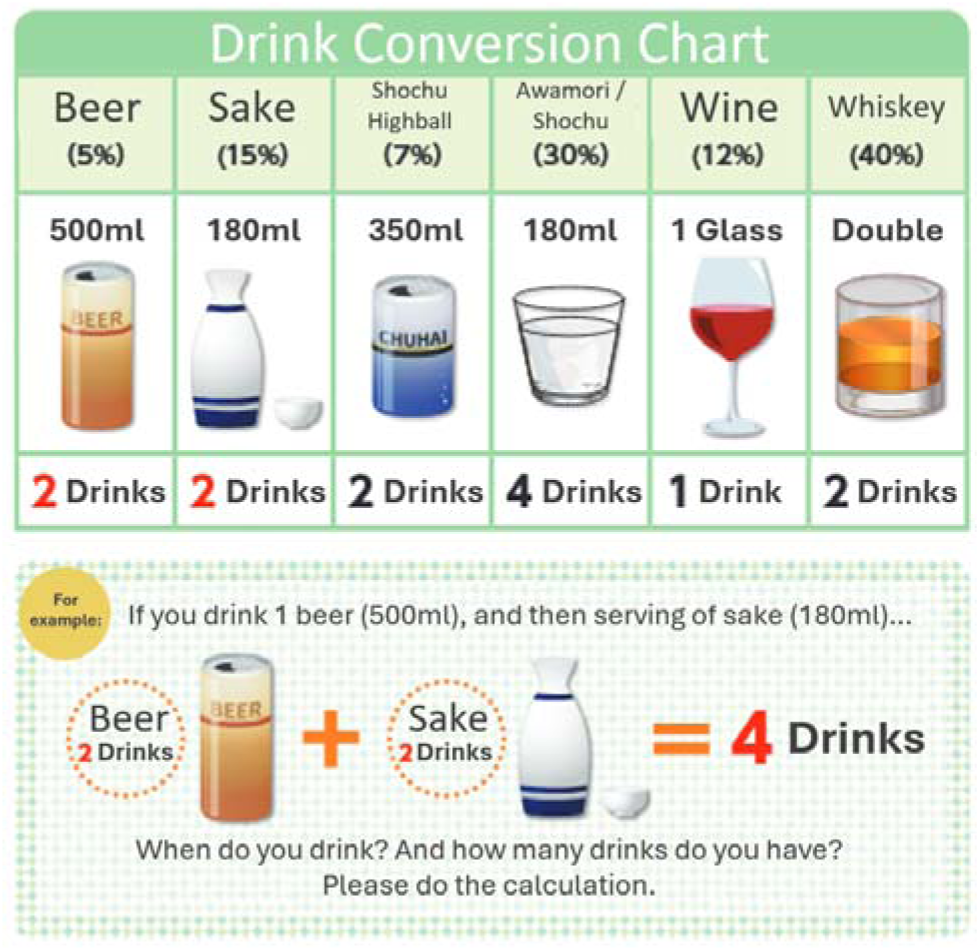
Standard drink conversion chart to estimate self-reported alcohol consumption with English translation. **Note**. Drink conversion chart is fair use and was created by the National Center for Addiction Prevention [28].

Probable *ALDH2*\*2 genotype will be screened through a brief two-question screener that asks about alcohol-related flushing, which has high sensitivity (90%) and specificity (88%) [29]. This screener asks: (a) “Do you have tendency to flush in the face immediately after drinking a glass of beer” (yes, no, or unknown), and (b) Did you have a tendency to flush in the face immediately after drinking a glass of beer during the first to second year after you started drinking (yes, no, or unknown)?” Answering yes to either question indicates probable possession of *ALDH2*2* and ALDH2 enzyme deficiency. Other screening questions include age, known pregnancy status, lifetime cancer diagnosis, and current alcohol use treatment.

### Assignment of Interventions

#### Randomization and Allocation Concealment

After informed consent is collected, participants will be randomized 1:1 and allocated to one of two groups immediately after screening, informed consent, and baseline assessment. The computer-generated randomization sequence will be stratified by number of standard drinks. Neo Marketing, a third-party panel survey organization, will use a centralized computer program to conduct randomization. Once randomized, allocation will immediately take place and the treatment or sham control intervention will commence. Because randomization and allocation are immediately conducted in a centralized computer, allocation concealment is guaranteed.

#### Blinding

The principal investigator will be blind to randomization and group allocation until after data analysis. Participants will not be blind to group allocation. However, because there is a sham intervention for the control, participants will be unaware if their treatment is the primary point of investigation. The public title of the study is, “Brief Intervention to Promote Healthy Behavior: Randomized Controlled Trial” as listed in the Informed Consent Document. Care providers are automated educational videos and technically blinded. All outcome measures are self-reported. Thus, the outcome assessor is the participant and blind to the study hypothesis. Data analysts will be blind to randomization and group allocation, as raw data will be coded upon delivery from the third-party panel survey organization.

### Interventions

#### BIGRE Group

BIGRE consists of an animated educational video with a host medical doctor avatar voiced by a professional voice-over artist. It provides spoken information (Multimedia Appendix 1, Table S1), animated content, and graphical depictions of graphs, charts, and genetic processes (Figure 3). The components will emphasize that a) the participant’s screening indicates that they are likely an *ALDH2*2* carrier, b) *ALDH2*2* carriers show increased risk for specific cancers that increases with increased alcohol consumption, c) the association of *ALDH2*2* with cancer risk is not deterministic, and d) non-or infrequent drinkers with *ALDH2*2* do not show elevated risk for alcohol-related cancers. Research on esophageal cancer risk associated with alcohol use in people with the *ALDH2*2* allele is wildly varied with relative risks (RR) ranging from 1.5 times the risk for all drinkers to 89 times the risk for heavy drinkers [10]. In an effort to avoid overstating risks, we will use very conservative estimates when explaining the risks to participants. Experimental event rate estimates for people with the *ALDH2*2* allele is based on estimates taken from the summary literature meant to demonstrate the exponential dose response with increasing alcohol consumption [10] and a control event rate of 1% [30]. Compared to non-drinkers, we estimated risk to be 2 times greater for light drinkers with the *ALDH2*2* allele, 2.4 times for moderate drinkers, and 4.4 times for heavy drinkers, noting that some research suggest the risks are much higher.

**Figure 3.**
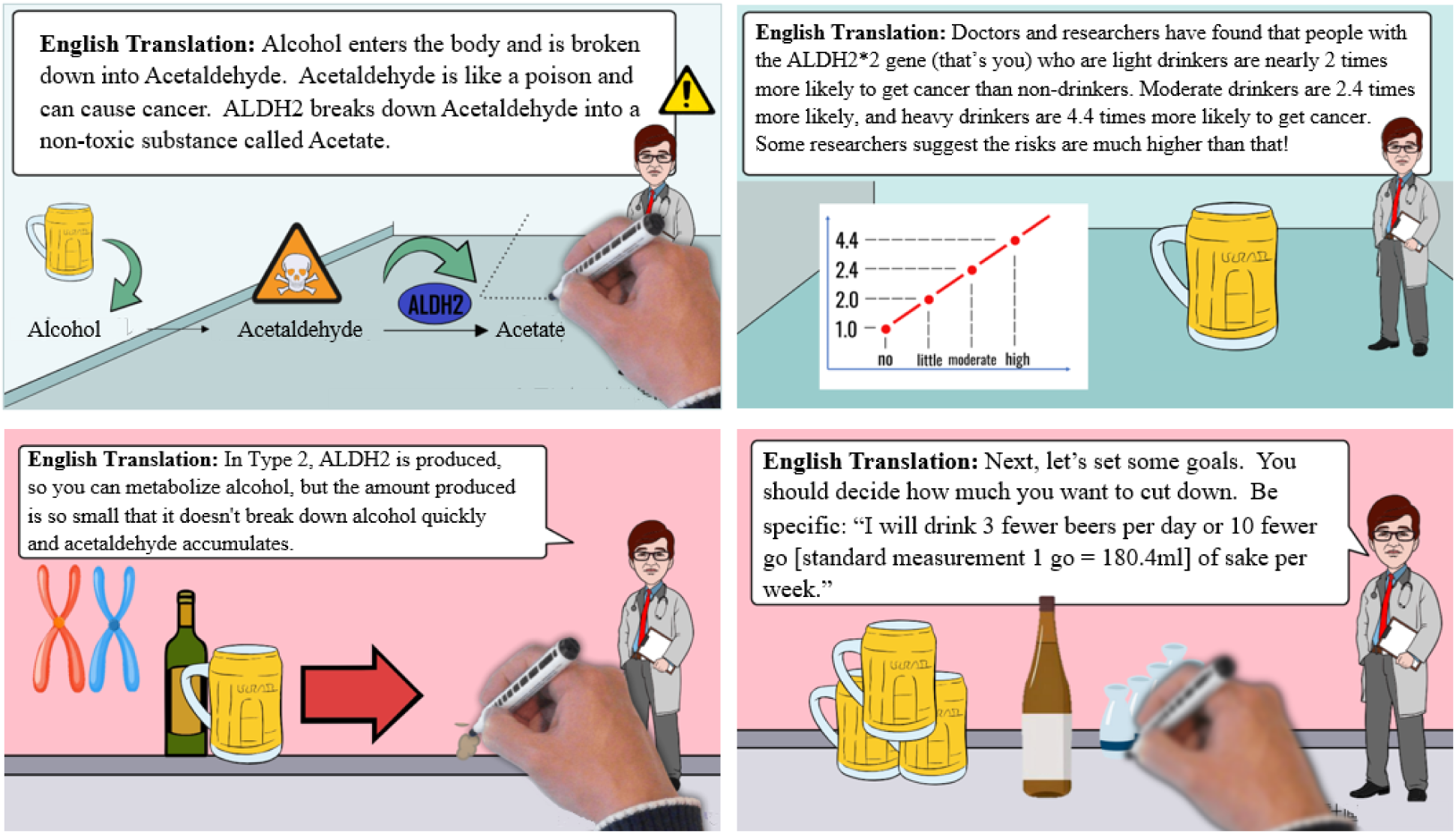
Unguided web-based brief intervention with genetic risk education (BIGRE) video screenshots with English translations. **Note**. All printed words are also spoken by a professional voiceover artist.

The video will last under 6 minutes (5:49) and incorporates the FRAMES approach [16,19–21]:

1. **Information:** Education on the ALDH2 enzyme, the influence of *ALDH2* genetic variations on alcohol metabolism, the relationship between possessing an *ALDH2*2* allele and esophageal cancer risk, and the risks of continued alcohol use.
2. **Feedback:** Informing the participant they are likely an *ALDH2*2* carrier
3. **Responsibility:** Stating that change is up to the participant to achieve the benefits of reducing alcohol use.
4. **Advice:** Providing pointers on how to reduce use using motivational enhancement techniques
5. **Menu of options**: Providing suggestions on how to develop a plan to reduce use and to start altering language to support sustain talk.
6. **Empathy:** Delivery of genetic risk education by supporting in an understanding tone and affirming that this change is not easy, even a small change is great.
7. **Supporting Self-Efficacy:** The narrator notes that decisions are up to the participant if they choose and provides options for self-help.

#### QoL Sham Control Group

The control group will receive a sham video that describes the importance of QoL and health, description of QoL concepts, ways to change health behaviors to improve QoL (Multimedia Appendix 1, Table S2). The control sham-intervention consists of an animated educational video with a host medical doctor avatar voiced by a professional voice-over artist. It provides spoken information, and animated content. The video will last just under 6 minutes (5:38).

### Measurements

Table 1 shows the plans for data collection at baseline and follow-up. Collection will be conducted through the Neo Marketing survey software. Data entry, coding, security, and storage will be managed by Neo Marketing software.

**Table 1.**
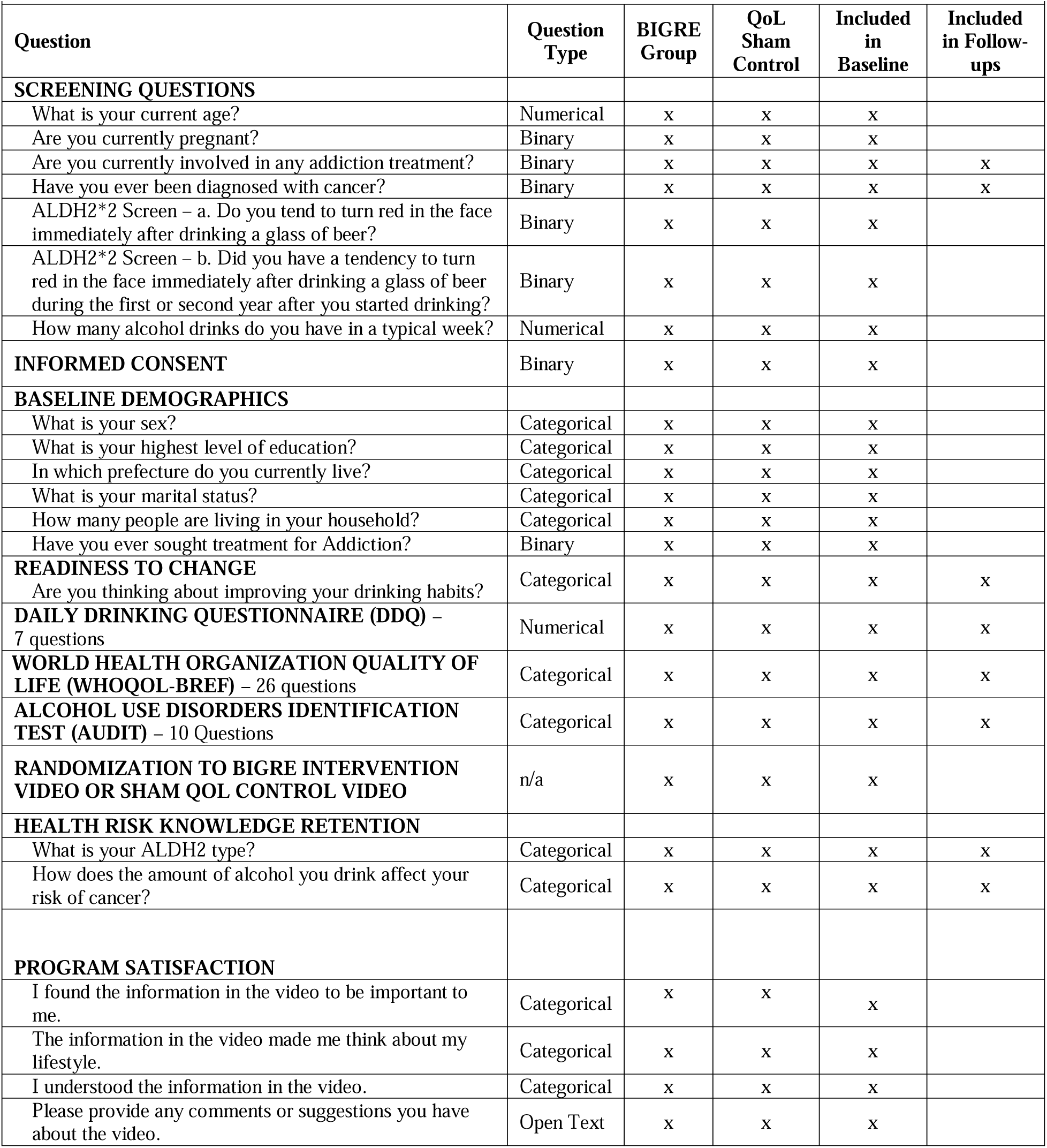
Plans for assessment and collection of data at baseline and follow-up assessments.

#### Primary Outcome

The primary outcome is mean endpoint past 4-week alcohol quantity at 3-months post-randomization. Alcohol quantity will be measured in number of standard drinks with the Daily Drinking Questionnaire (DDQ) [31]. Participants will be provided with a definition of a standard drink. Standard drinks are approximately: 250ml beer (∼5% alcohol), 90ml wine/sake (∼15% alcohol), 45ml whiskey shot (40% alcohol). The DDQ asks, “Consider a typical week during the last 3 months. How much alcohol, on average (measured in number of drinks), do you drink on each day of a typical week?” The participants will then answer seven items representing each day of the week. For example, “On a typical Monday, I have drinks.” For each day, participants enter a numerical response representing the number of alcoholic beverages consumed on an average day. The responses for a typical week are then summed and averaged over the past 30 days. The DDQ has been used in Asian populations with known *ALDH2*2* alleles [23] and has demonstrated good internal validity (Cronbach’s α = 0.83) [32] adequate test-retest reliability [33] and criterion validity [34].

#### Secondary Outcomes

Secondary outcomes include the quantity in standard drinks at 1- and 2-month follow-ups; alcohol use in grams, improvement, severity, motivation to change, health knowledge retention, and quality of life at 1-, 2-month, and 3-month follow-ups post-randomization. Participant satisfaction will be measured after the intervention program has been delivered.

#### Quantity

Mean endpoint past 4-week alcohol use quantity will also be measured at 1- and 2-months post-randomization in standard drinks and in grams. Alcohol quantity in grams will alternatively be converted from DDQ with participants citing what they typically drink (340ml beer, 100ml wine/sake, or 250ml highball/Collins. Alcohol use quantity measured as within group change will also compare pre- and post-intervention improvement.

#### Severity

Alcohol use severity will be assessed with the alcohol use disorders identification test (AUDIT) Japanese version at all timepoints. The AUDIT is a 10-item screening instrument scored 0-40, rating alcohol misuse from no/low-risk to severe [35]. The AUDIT has been validated with multiple international samples with high sensitivity (0.90s) and specificity (0.80s) with a cutoff of 7 [35], strong test-retest reliability (K-S Test = 0.80s) [36], and moderate stability for severity level screening (ICC = 0.56, 95% CI = 0.52-0.60) [37], Internal consistency of the Japanese version of the AUDIT is considered satisfactory (Cronbach’s α = .67) [38].

#### Readiness to change

Readiness to change alcohol use will be measured at all timepoints using questions from the Japanese National Health and Nutrition Survey [39], which include levels of: already working on improvement (maintenance/action), intending to improve (contemplation/planning), no intention to improve (precontemplation), and no improvement needed (precontemplation).

#### Quality of life

QoL will be measured at all timepoints with the Japanese version of the WHOQOL-BREF [40], a 26-item measure assessing QoL in 4 domains of physical health, psychological, social, and environmental. With reasonable internal consistency reported by domain (Cronbach’s α = 0.68-0.82), and good discriminant validity between well and sick samples (*p*s < 0.01). For the Japanese sample, internal consistency ranges from 0.66-0.75.

#### Health risk knowledge retention

Health risk knowledge retention will be assessed at all timepoints with a single question asking, “Do you recall your potential alcohol risk genotype?”

#### Program satisfaction

Participant satisfaction will only be measured after intervention completion for the BIGRE group with 4 survey statements (5-points, Strongly Disagree to Strongly Agree) reporting, “I found the program information to be important”, “The program information made me think about my lifestyle.”, and “I understood the content in the program information”. Finally, qualitative comments will be collected that simply prompt participants to, “Please provide any comments or suggestions you have about the program content.”

### Monitoring

A data monitoring committee will not be used due to very low risks, self-reported endpoint data selection, intervention brevity, and general population selection [41]. In the rare event of an adverse event occurring during the brief educational intervention, the program would be completed before the ability to stop the program could be determined by an outside monitor. Thus, safety data will not be collected, and participants will be informed that they may stop and withdraw from the trial at any time they may feel uncomfortable.

#### Anticipated Risks and Benefits for Participants

The interventions will consist of a screening (2 minutes), survey (15-20 minutes) and brief intervention (approximately 6 minutes) providing education on probable genetic information identified as an alcohol-related cancer risk. Thus, the intervention will therefore be considered as minimally invasive with no serious health risks expected. There are always possible psychological and time burdens associated with participation of health education and responding to questionnaires. Conversely, the only comparable study investigating alcohol use, *ALDH2*2*, and cancer risk reported no adverse events and high participant satisfaction, interest, and engagement [23]. The risks and benefits will be fully disclosed and explained to potential participants via informed consent. Neo Marketing would manage any participant complaints in line with their reporting procedures. Of which, participants agree to standard participant and private policy agreements.

#### Remuneration

Participants will be compensated with Neo Marketing points that can be redeemed in various ways through the company website. Remuneration is aligned with Neo Marketing standards. Points are valued at ¥700 JPY for completing the screening, baseline assessment, and intervention. Participants will be compensated with points valued at ¥200 for completing each of the three follow-up assessments. Those who finish all assessments will receive a completion bonus of ¥1,000. In total, a final remuneration of ¥2,300 (∼$16.00 USD) can be awarded in point equivalents.

#### Methods to Decrease Protocol Deviations

In this study, a one-time BI will be provided to study participants immediately upon study enrollment. The intervention will be administered through a fully automated computer program, making deviations only due to error in connectivity. Because the intervention consists of a short video, it is unlikely that the actual intervention differs from the method described in the protocol. After the intervention and during the study follow-up phase (3 months post-randomization), participants have no restrictions for concomitant care or co-interventions to reduce alcohol consumption, which is measured at each follow-up. Treatment cannot be discontinued or modified after the completion of the BI (5:49) and this intervention was deemed to be of low risk as it is a general population and of an educational nature. However, participants may request to be removed from the study at any time and for any reason.

### Ethical Considerations

The present study has been approved by the Kyoto University Graduate School of Medicine Ethics Committee (C1711-1) on July 9, 2025. All participants will provide e-consent. If protocol amendments are required after finalization, changes will be documented in the appropriate medium: ethics review board amendments, trial registry, or results publications. Informed consent will be collected by Neo Marketing through a web portal. Participants will be provided with an explanation approved by the Kyoto University Ethics Committee. Multimedia Appendix 1, Table S3 provides the informed consent information addressing confidentiality, handling of personal information, access to data, data storage and disposal procedures, and plans for dissemination of results.

### Statistical Analyses

#### Sample Size

Sample size analysis suggests 64 participants (32 per group) would give 80% power (α = 0.05, β = 0.2) to detect a standardized mean difference (SMD) of 0.7. Previous findings indicate an expected 50% reduction for the experimental condition [23]. Our proposed inclusion criteria requires a minimum of 14 standard drinks in the past 30 days. Thus, for our initial study a 7-drink difference is expected from the minimum drinkers, and we will assume large variability from a general population (SD=10 drinks). Accounting for attrition, the available budget, multiple frequent follow-ups, and analytic methods, the target sample will be 100 participants (50 per group), giving 80% power to detect and SMD of 0.5.

#### Analyses

The full analysis set (FAS) will include the largest population for analysis based on the intention-to-treat (ITT) principle, excluding participants who: Withdraw consent and disallow the use of their data after enrollment or have no efficacy-related data available. The safety analysis set (SAS) will include all randomized participants, including those who discontinue or drop out of the study. See the full statistical analysis plan (SAP) for further information (Multimedia Appendix 1, Table S4)

#### Primary Analysis

The primary analysis will use the FAS population and assess the primary outcome (alcohol use quantity with the DDS at 3 months follow-up) with a generalized linear mixed model (GLMM) assuming a negative binomial distribution to address possible overdispersion in the count data [42]. In this model, the treatment group, time point (months 1, 2, and 3), and their interaction will be modeled as fixed effects, and random effects include both the random intercept and the slope for time account for individual variability in baseline outcome levels and temporal trends. The model will provide the estimated marginal means (EMMs) and their 95% confidence intervals (CIs) at 3-months.

A sensitivity analysis using the SAS population will be conducted for the primary outcome with only complete follow-up cases included. We will conduct subgroup analyses for the primary outcome to explore potential interactions between the group and the following pre-specified factors: age, sex, severity, health knowledge retention. These factors will be included as covariates in the model, along with their interaction terms with the intervention group. The p-value for each interaction term will be estimated.

#### Secondary Analyses

Binary secondary outcomes such as health knowledge retention will be analyzed using a GLMM with a binomial distribution. The model specification will follow the same structure as that used for the primary outcome analysis. For continuous secondary outcomes such as alcohol use in grams, readiness to change, and QoL, we will use mixed model for repeated measures (MMRM). These models will include intervention group, time points (months 1, 2, and 3), and the interaction between intervention group and time as covariates. If the sample size is too small to achieve convergence, simpler models will be applied.

#### Reporting

In both GLMM and MMRM models missing data are assumed to be “missing at random” (MAR), and analyses incorporate all available repeated measures from all participants and account for within-subject correlations over time. Although missing values are not directly imputed, the observed data contribute to estimating parameters that implicitly inform inference in the presence of missing data. Secondary outcomes including participant satisfaction and safety will be summarized descriptively.

Alpha will be set a *p* < 0.05. Effect sizes will be represented with a standardized mean difference (SMD) using baseline SDs for continuous outcomes, and odds ratio, risk difference and number needed to treat (NNT) for binary outcomes. All analyses will be conducted using RStudio version 2024.12.0 Build 467.

## EXPECTED RESULTS

If efficacious, a simple, brief, and inexpensive intervention to reduce unhealthy alcohol use could be an attractive addition to public health initiatives and primary care clinics. For the primary outcome, it is expected that the experimental BI condition will significantly reduce alcohol consumption greater than the control condition and information about potential subgroup differences between severity levels, health risk knowledge retention levels, sex, and age will provide information about future interventions of this nature. Further groups differences are expected from secondary analyses.

The proposed research project adds original contributions in many ways. To our knowledge, this would be the first study in Japan to investigate the efficacy of an unguided web-based BI for alcohol use. Second, the proposed research would improve upon previous studies by considerably reducing treatment costs with the elimination of genotyping, making the intervention non-invasive. This could also improve feasibility by substituting a short screener that identifies probable *ALDH2*2* genotype. Third, BI is well studied in the U.S. and Europe.

However, little is known about the efficacy in Asian countries with different drinking cultures, consequences, and effective motivational factors for moderated drinking. Fourth, the existing *ALDH2* BI studies were done with university students, most of whom drank alcohol at low levels. The proposed research will improve upon the existing U.S. study by recruiting from the general population who report moderate to high alcohol consumption.

## Data Availability

All deidentified data produced in the present study will be made available upon reasonable request to the authors.

## AUTHOR DECLARATIONS

### Author Approval

All authors have seen and approved the manuscript.

## Acknowledgements

The authors would like to thank Kaori Okazaki and Sei Shirai for their work on the intervention videos. No AI was used in any part of this manuscript’s creation.

## Funding

This study was funded by a grant (24K20239) from the Japan Society for the Promotion of Science.

## Data Availability

Deidentified data can be made available upon reasonable request to the authors.

## Competing of Interests

TAF reports personal fees from Boehringer-Ingelheim, Daiichi Sankyo, DT Axis, Micron, Shionogi, SONY and UpToDate, and a grant from DT Axis and Shionogi, outside the submitted work; In addition, TAF has a patent 7448125 and a pending patent 2022-082495, and has licensed intellectual properties for Kokoro-app to DT Axis.MS is employed in the Department of Neurodevelopmental Disorders, Nagoya City University Graduate School of Medical Sciences, which is an endowment department supported by the City of Nagoya 7448125 and a pending patent 2022-082495, has received a personal fee from SONY outside the submitted work, and has licensed intellectual properties for Kokoro-app to DT Axis. RS is employed by CureApp, Inc. He received grants from the Osake-no-Kagaku Foundation and the Mental Health Okamoto Memorial Foundation. He holds multiple pending patents (JP2022049590A, US20220084673A1, JP2022178215A, JP2022070086, JP2023074128A). All the other authors report no competing interest.

## Authors Contributions

ES conceived the study. ES, TW, SK, MS, and TAF provided substantial contribution to the design of the study during its development. YK, MTS, SS contributed to the development of the procedures. ES and YL contributed to the statistical methods. RS and TAF provided supervision to the entire trial design. All authors revised the study protocol critically for important intellectual content and contributed to its improvement. All authors gave the final approval to publish the current protocol and agreed to be accountable for all aspects of the work in ensuring that questions related to the accuracy or integrity of any part of the work are appropriately investigated and resolved.

## Abbreviations

ALDH2*2: aldehyde dehydrogenase 2*2 allele
BI: brief interventions
FRAMES: feedback, responsibility to change, advice, menu of treatment options, empathy, and supporting of self-efficac
BIGRE: brief intervention using genetic risk education
QoL: quality of life
DDQ: Daily Drinking Questionnaire
AUDIT: alcohol use disorders identification test
WHOQOL-BREF: World Health Organization quality of life – brief measurement
SAP: statistical analysis plan
SMD: standardized mean difference (Cohen’s d)
FAS: full analysis set
ITT: intention-to-treat
SAS: safety analysis set
GLMM: generalized linear mixed model
EMMs: estimated marginal means
MMRM: mixed model for repeated measures
NNT: number needed to treat

## MULTIMEDIA APPENDIX

**Table S1.**
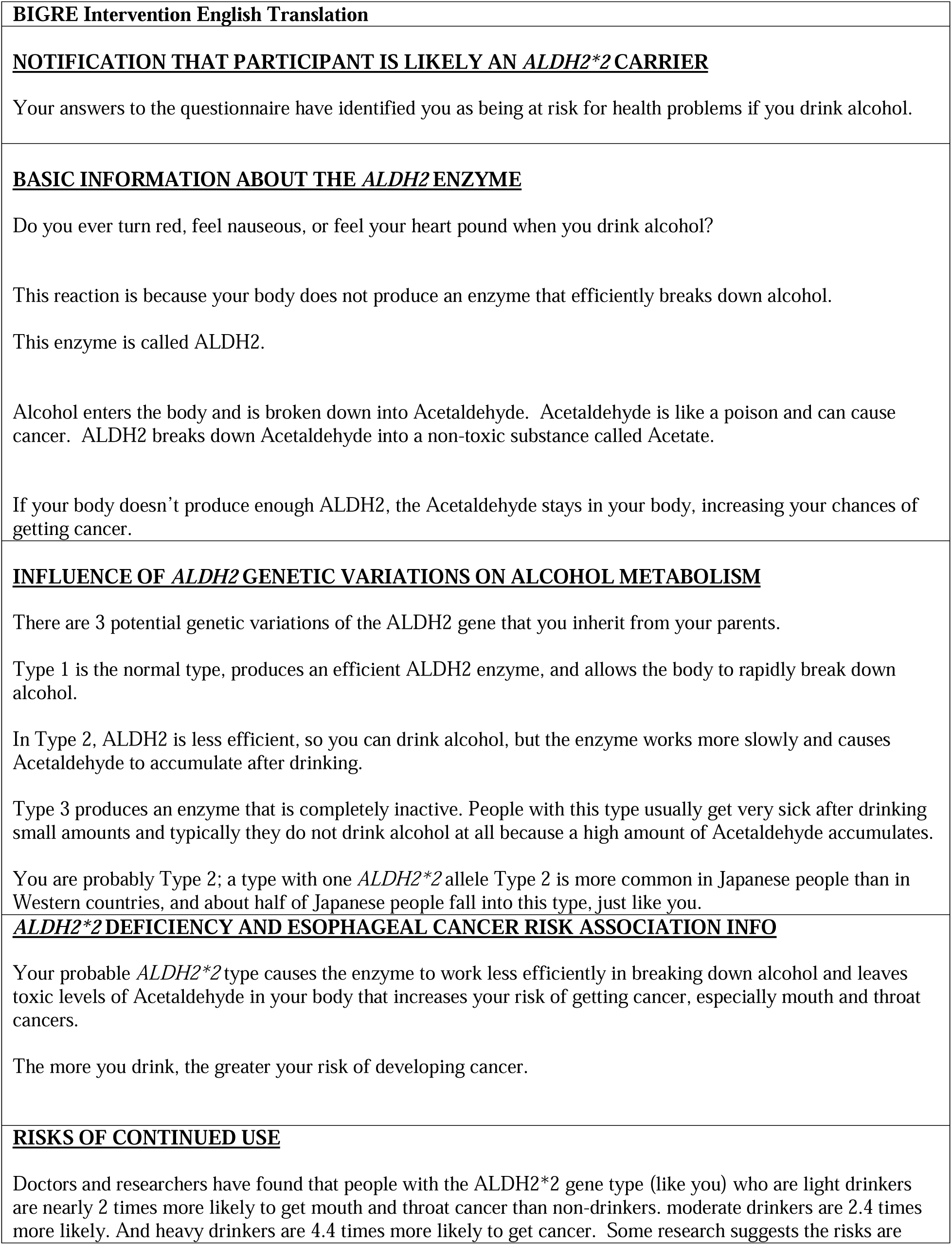
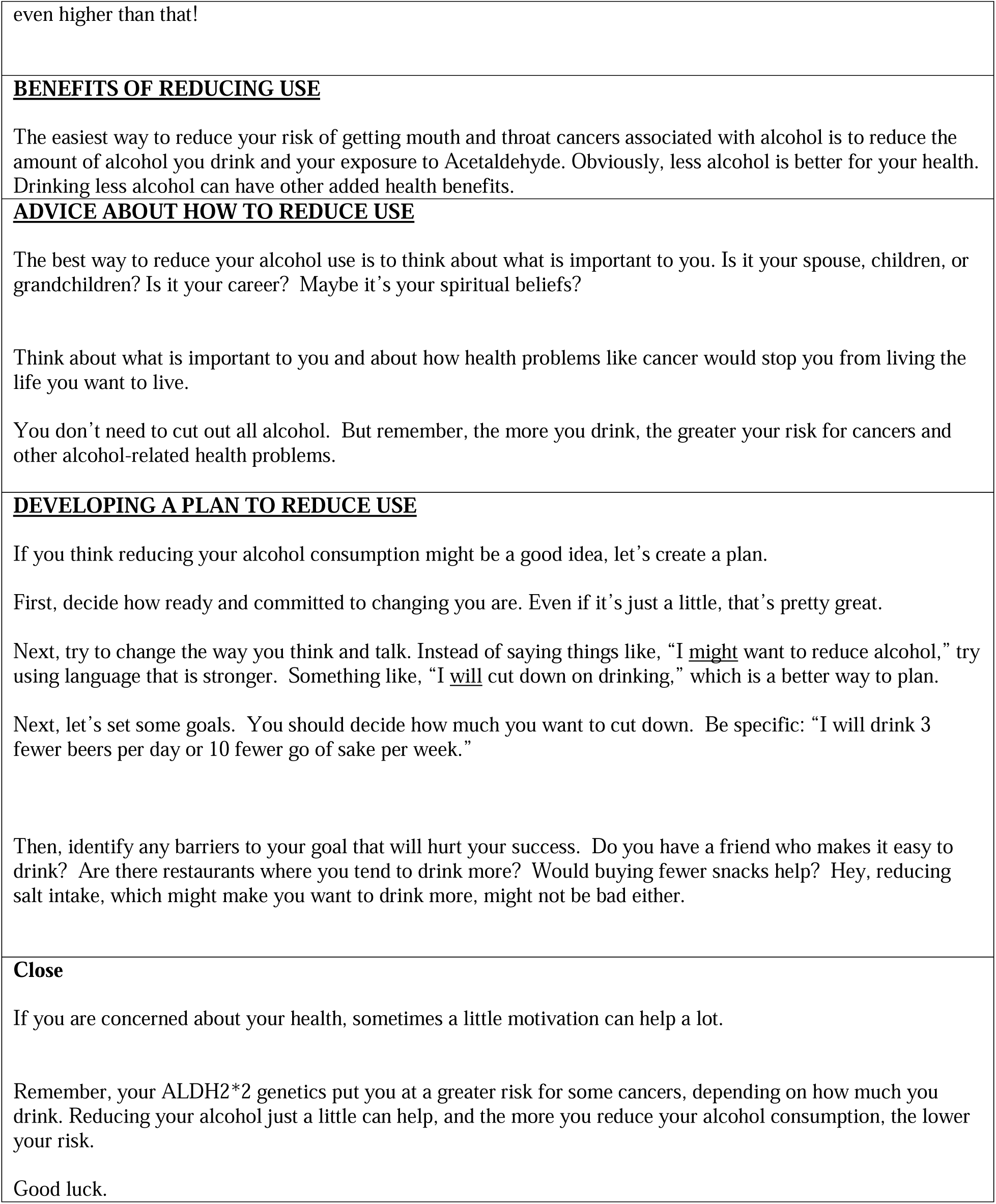
Brief Intervention with Genetic Risk Education (BIGRE) video intervention script.

**Table S2.** Quality of Life (QoL) Sham Control video script.

| **QoL Sham Control English Translation** |
| --- |
| Many people can benefit from improving their quality of life. |
| Have you been having trouble sleeping at night or feeling depressed or anxious lately?<br>This can be associated with a decreased quality of life.<br>Furthermore, it has been shown that a lower quality of life can lead to lower efficiency in learning and working. |
| Quality of life is assessed by the WHOQOL-BREF, which measures four domains with a 26-item scale.<br>The first domain is physical health. It is related to the presence or absence of illness, physical pain, or discomfort.<br>The second domain is mental state. It involves whether you enjoy life, accept your appearance, and are satisfied with your ability to do your job.<br>The third domain is social. It concerns whether you are satisfied with your relationships, your sex life, and the support of your friends.<br>The fourth domain is environmental. It is related to your accessibility to medical facilities, social services, and whether you are satisfied with your surrounding transportation system. |
| A better quality of life means a healthier and more satisfied body and mind. Plus, quality of life gives you a greater ability to enjoy life. |
| Lower quality of life can lead to sleep disturbances, depressed mood, anxiety, and decreased work productivity. |
| There are countless ways to improve quality of life.<br>For example, you can take a long bath, take deep breaths, go out, or meet with other people.<br>For example, you can reflect on what you accomplished today, plan your life, or watch people.<br>For example, you can take an active role in community activities, take up a new sport, or help others.<br>You may also want to just sit and think about things or practice mindful contemplation. |
| The best way to change something to improve the quality of life is to think about what is important to you. Is it your spouse, children, or grandchildren? Is it your friends? Is it your career? Maybe it's your spiritual beliefs? Think about what is important to you and about how health and mental problems would stop you from living the life you want to live.<br>You don't need to make a whole change. But remember, if you do not make changes at all, you yourself will not change at all. |
| If you think doing something to improve the quality of life might be a good idea, let's create a plan.<br>First, decide how ready and committed to changing you are. Even if it's just a little, that's pretty great.<br>Next, try to change the way you think and talk. Instead of saying things like, "I <u>might</u> do **," try using language that is stronger. Something like, "I <u>will</u> **," is a better way to plan.<br>Next, let's set some goals. You should decide how much you want to make changes. Be specific: "I will take a bath for 30 minutes, take a deep breath once a day, or go on a gym twice a week."<br>Then, identifying barriers to your goal will help you be successful. Does skipping breakfast prevent you from being active? Is the cost of joining a sport club an obstacle? Would searching sports you can do near your house help? Hey, thinking of doing something at home might not be bad either. |
| You don't have to do it for others. You can do what you enjoy doing. |
| Just try a few new things, it can really help improve your quality of life. |

**Table S3.**
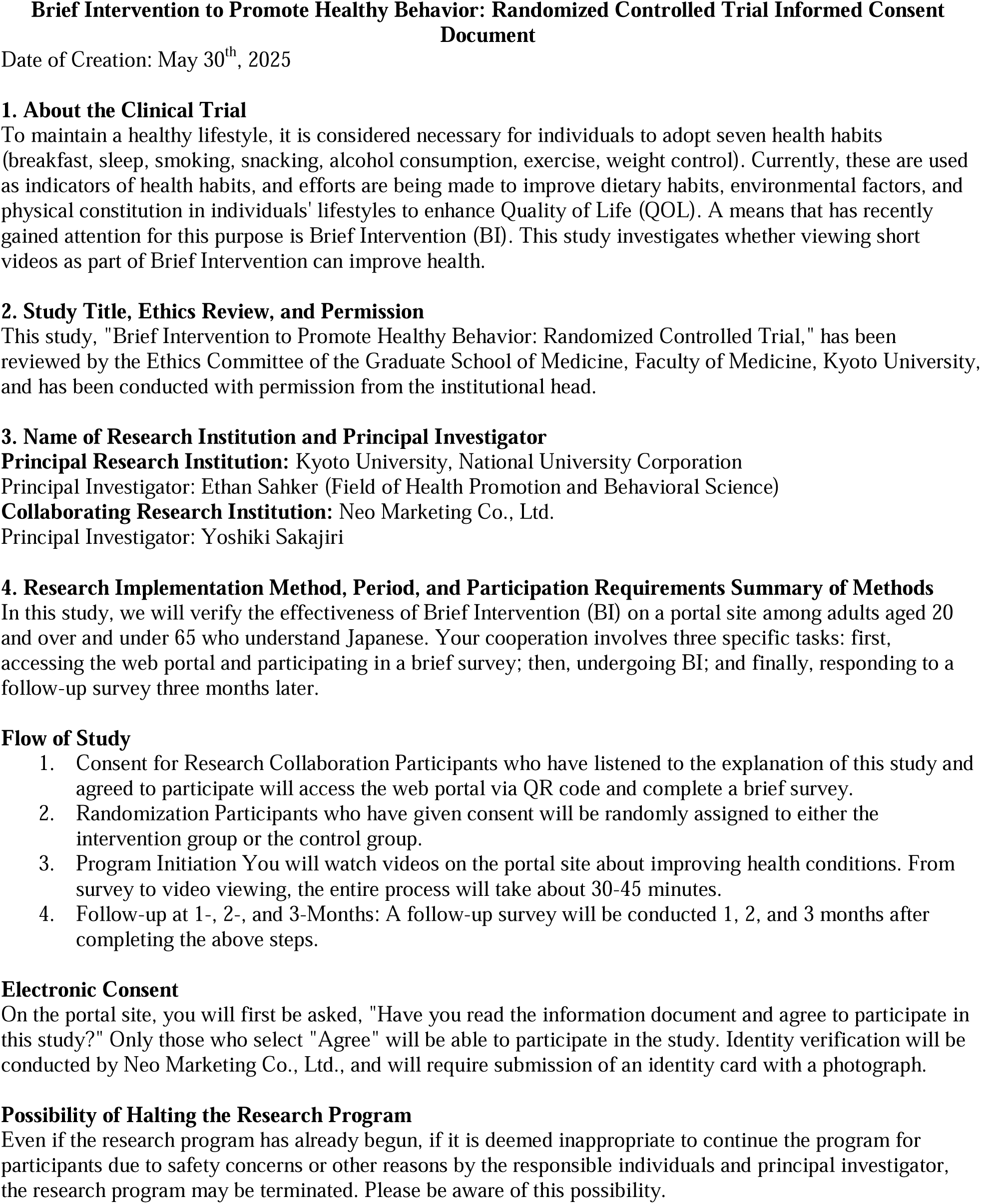
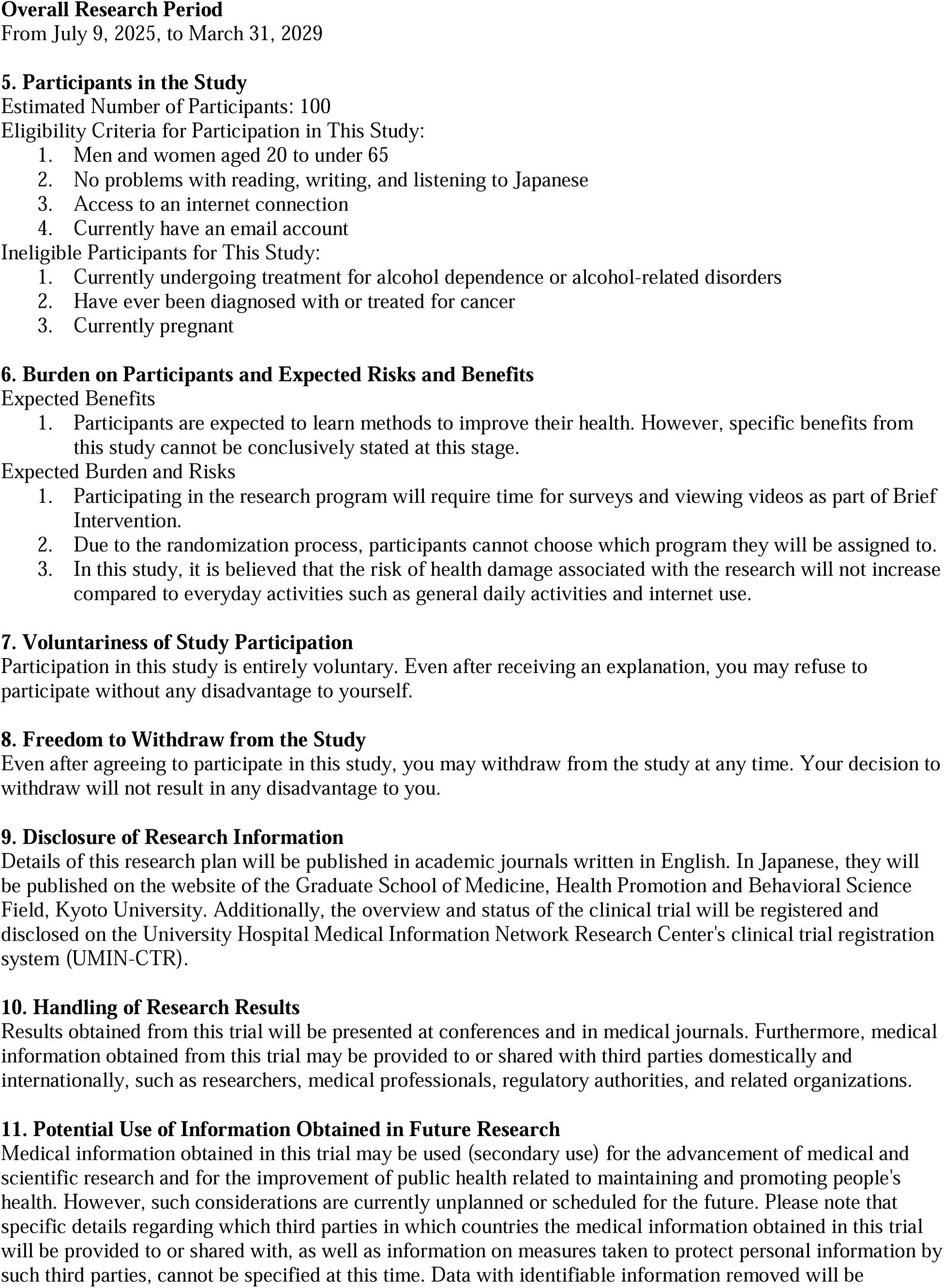
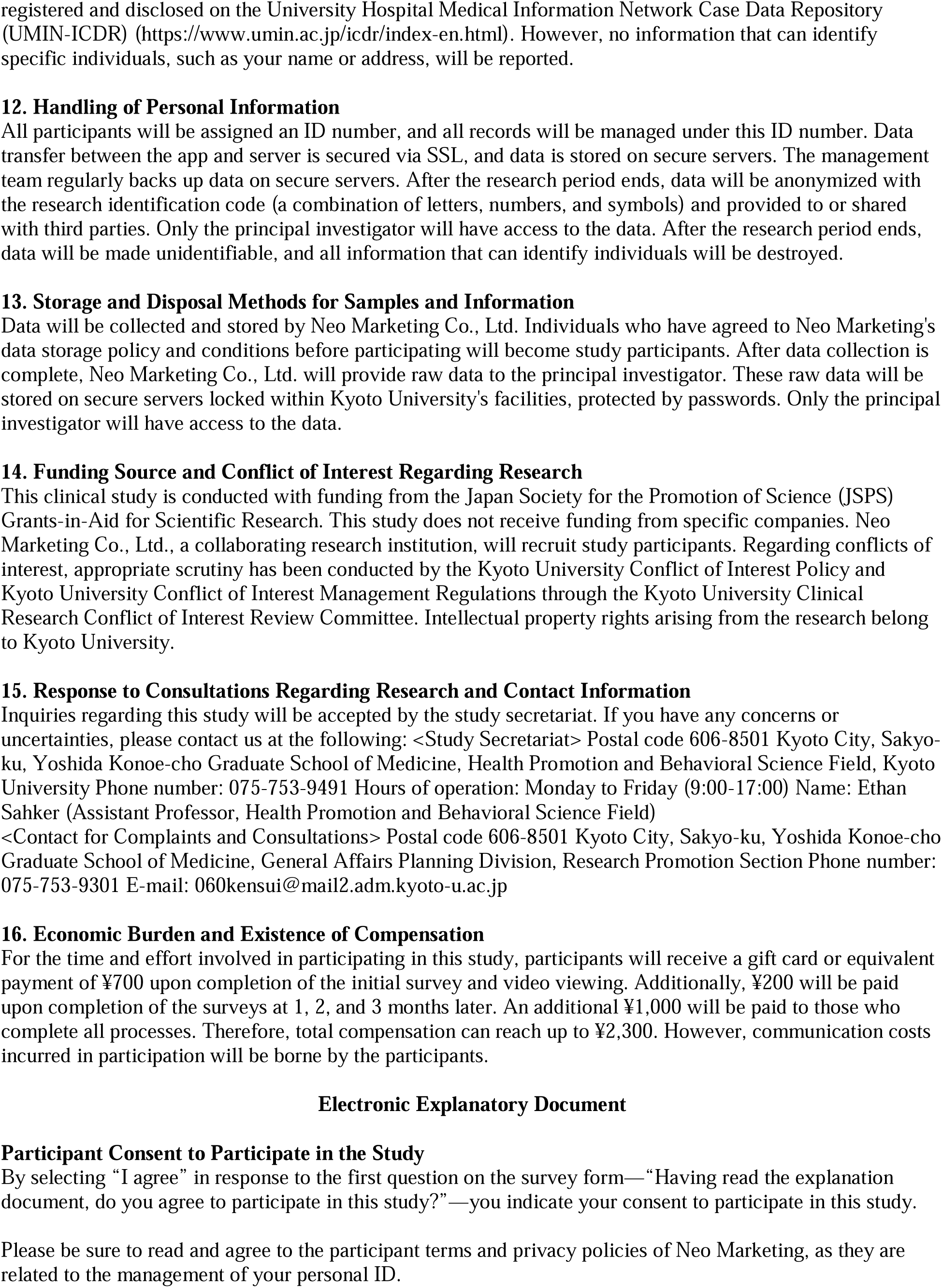
Informed consent document (translated from the original Japanese)

**Table S4.**
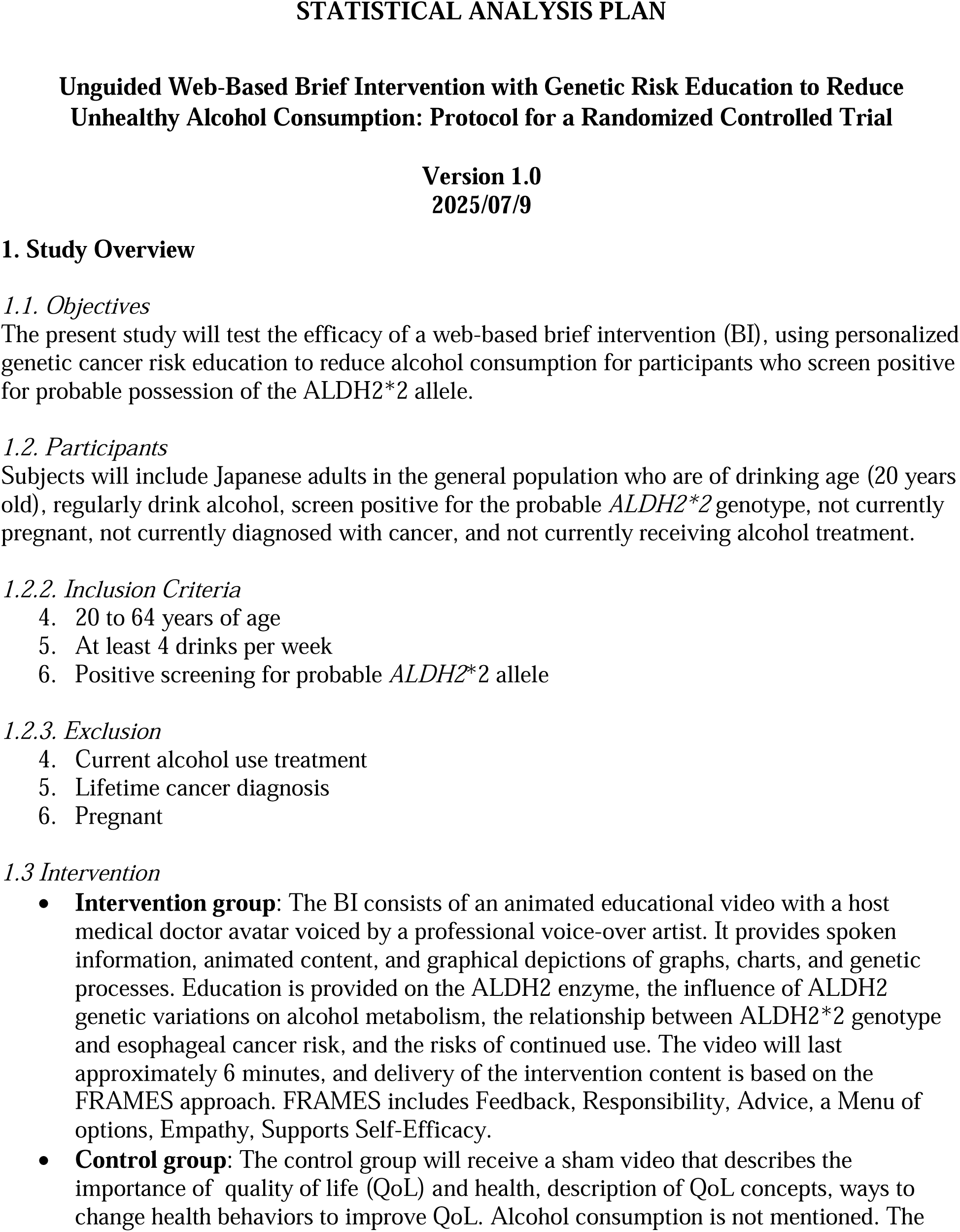
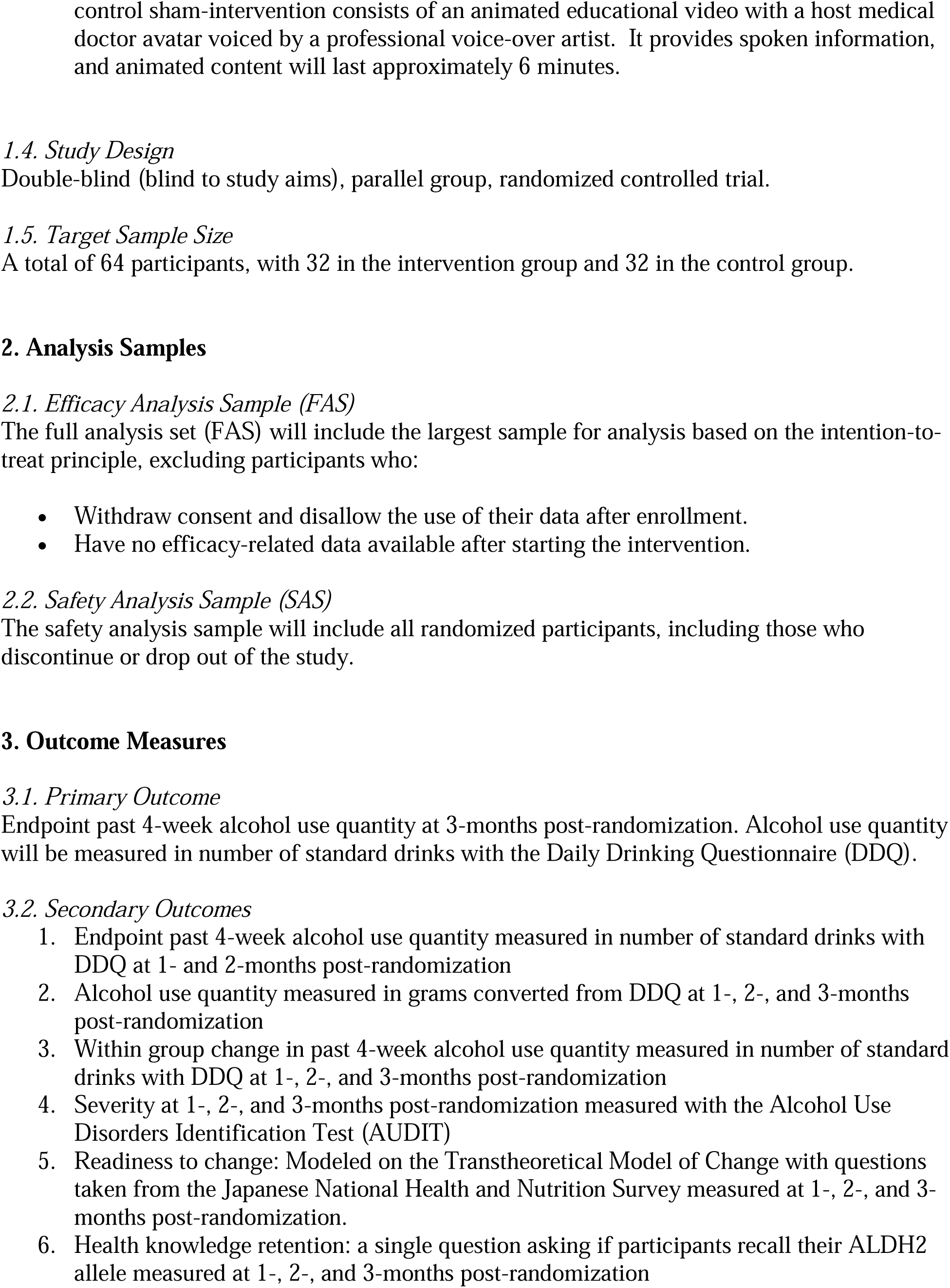
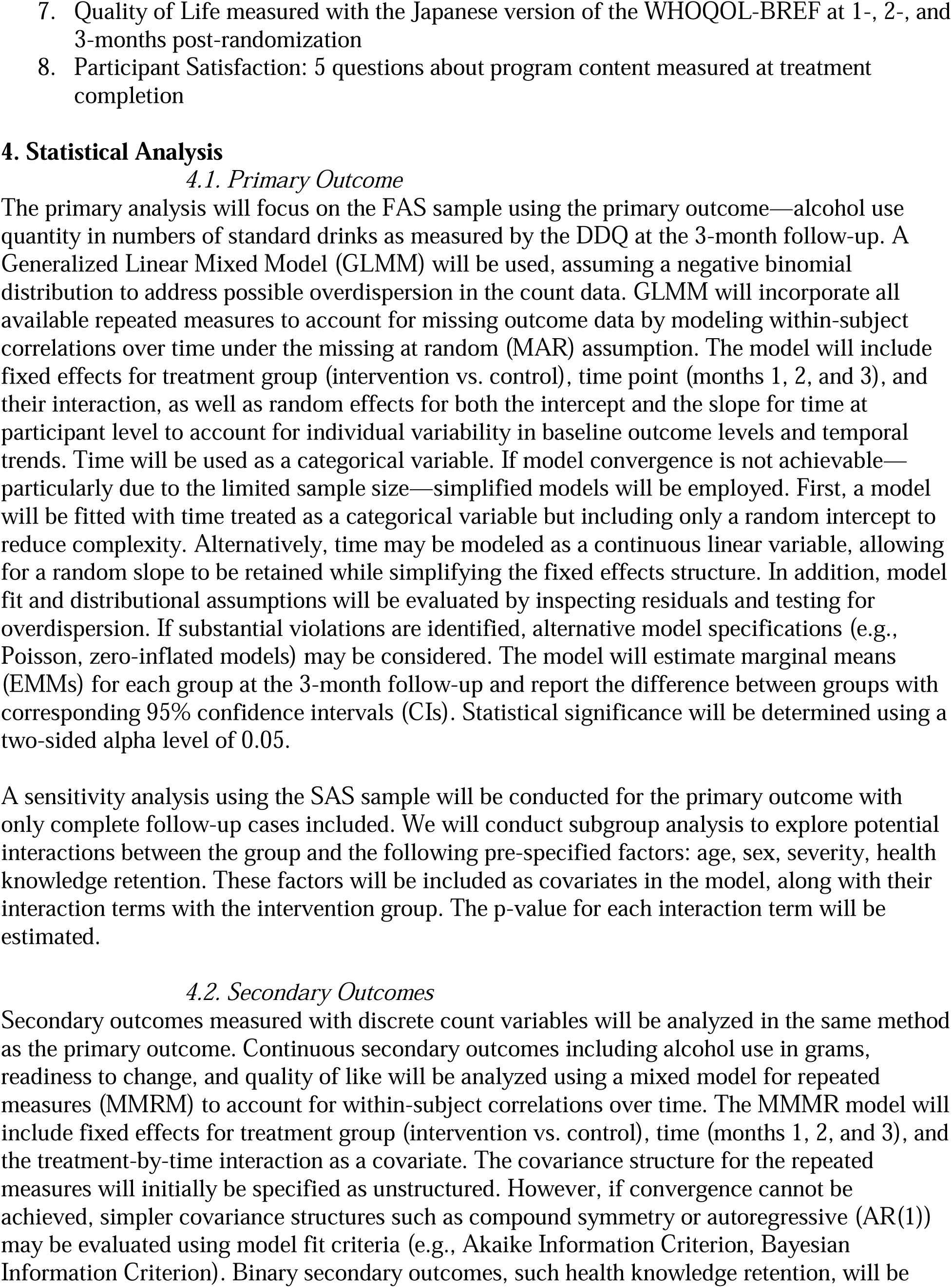
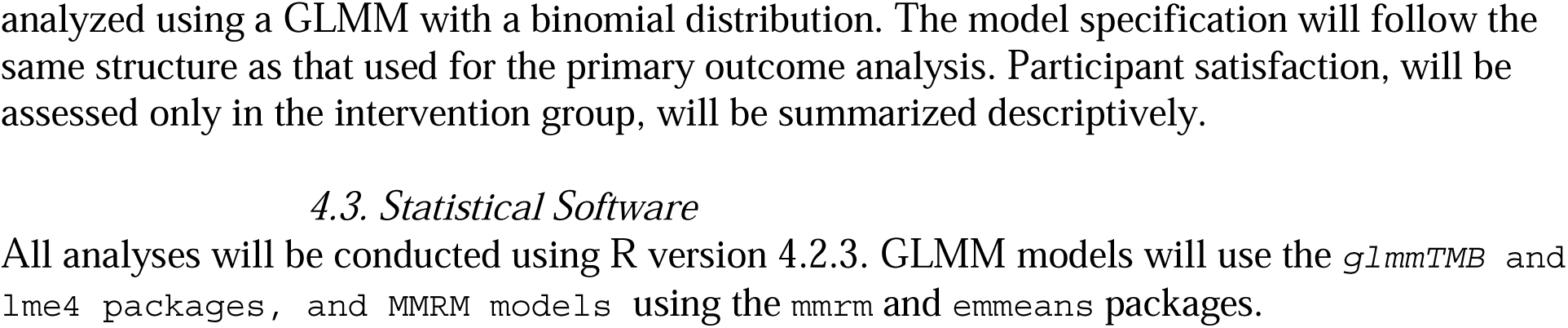
Statistical Analysis Plan.

## Notes

### Clinical Trial

UMIN000058012

### Author Declarations

Ethics committee of Kyoto University Graduate School of Medicine gave ethical approval for this work (C1711-1).

